# Effect of face-covering use on adherence to other COVID-19 protective behaviours: a systematic review

**DOI:** 10.1101/2023.04.11.23288200

**Authors:** Adam Millest, Sidra Saeed, Charles Symons, Holly Carter

**Affiliations:** Behavioural Science and Insights Unit (BSIU), UKHSA, London, United Kingdom

## Abstract

During the COVID-19 pandemic, concerns were raised that face covering use may elicit risk compensation; a false sense of security resulting in reduced adherence to other protective behaviours such as physical distancing. This systematic review aimed to investigate the effect of face covering use on adherence to other COVID-19 related protective behaviours. Medline, Embase, PsychInfo, EmCare, medRxiv preprints, Research Square and WHO COVID-19 Research Database were searched. All primary research studies published from 1 January 2020 to 17^th^ May 2022 which investigated the effect of face covering use on adherence to other protective behaviours in public settings during the COVID-19 pandemic were included. Papers were selected and screened in accordance with the PRISMA framework. Backwards and forwards citation searches of included papers were also conducted on 16^th^ September 2022, with eligible papers published between 1^st^ January 2020 and that date being included. A quality appraisal including risk of bias was assessed using the Academy of Nutrition and Dietetics’ Quality Criteria Checklist. This review is registered on PROSPERO, number CRD42022331961. 47 papers were included, with quality ranging from low to high. These papers investigated the effects of face covering use and face covering policies on adherence to six categories of behaviour: physical distancing; mobility; face-touching; hand hygiene; close contacts; and generalised protective behaviour. Results reveal no consistent evidence for or against risk compensation, with findings varying according to behaviour and across study types. There is a suggestion that face covering use might reduce face-touching and face covering mandates might increase mobility, though the lack of robust evidence means these are tentative claims. Evidence on the other protective behaviours is largely inconsistent, and therefore confident conclusions cannot be made in these areas. Any policy decisions related to face coverings must consider the inconsistencies and caveats in this evidence base.

## 1. Introduction

The COVID-19 pandemic highlighted the important role that public behaviour can play in the control of infectious disease. Prior to the introduction of vaccination, Prior to the introduction of vaccination, measures to control the spread of COVID-19 consisted primarily of non-pharmaceutical interventions (NPIs), e.g. hand cleaning, reducing contact with others, and wearing a face covering to effectively reduce the spread and mortality rate of COVID-19 (1). The extent to which populations effectively undertook these behaviours was therefore crucial to controlling the spread of COVID-19.

A key behaviour which was used to control the spread of COVID-19 was the use of face coverings, with over 160 governments mandating or recommending the use of face coverings during the COVID-19 pandemic (2). Many previous systematic reviews have been carried out on the transmission reduction efficacy of face coverings and other NPIs (e.g.,3, 4). This review aims to explore how face-covering use impacts on and interacts with other COVID-19 protective behaviours.

At the onset of the pandemic, concerns were raised that the use of face coverings may provide a false sense of security, leading to lower adherence to other protective measures such as hand hygiene and physical distancing; a phenomenon known as ‘risk compensation’ (5). Arguments exist both for and against the existence of risk compensation (6, 7), and one review was carried out early in the pandemic to examine the extent to which risk compensation may affect public behaviour (8). However, given the paucity of research into COVID-19 specifically, this review necessarily relied on research into risk compensation in other contexts.

More recently, the concern about the potential impact of face coverings on other protective behaviours has prompted several studies examining this in the context of COVID-19, and there is now a substantial body of research that can contribute to discussions around risk compensation in this context. Given the widespread recommendation and mandation of face coverings during the COVID-19 pandemic, it is essential to consider all available evidence in order to understand the potential impact of face covering use on adherence to other types of protective behaviours in the context of COVID-19.

The aims of this systematic review were therefore to: identify and summarise the findings from studies examining the effect of face covering use on other protective behaviours in the context of COVID-19; assess available evidence relating to the effect of face covering use on adherence to other COVID-19 protective behaviours. This aim was achieved and findings will help to inform decisions regarding if and when it is appropriate to recommend or mandate face covering use during future infectious disease outbreaks.

## 2. Methods

Details of the the protocol for this systematic review were registered on PROSPERO before screening took place. The protocol’s number is CRD42022331961 and can be accessed at https://www.crd.york.ac.uk/PROSPERO/display_record.php?RecordID=331961 (9). As can be seen in the online record, the protocol was amended on 20^th^ May 2022; these amendmends comprised edits to address inconsistencies with the regards to the decision to use the term ‘face-covering’ or ‘face mask’ throughout the protocol. Further amendments were made on 5^th^ September 2022, 21^st^ September 2022, 1^st^ November 2022, and 3^rd^ January 2023; these comprised updating the estimated completion date and also updating which stages of the project had been completed (e.g., data extraction, data analysis).

### 2.1 Eligibility criteria

Studies were eligible if they reported primary quantitative or qualitative research relating to the impact of face covering use on adherence to other protective behaviours during the COVID-19 pandemic. Protective behaviours of interest included but were not limited to: COVID-19 physical/social distancing, hygiene (e.g., avoidance of face-touching, regular handwashing), staying at home, and reducing travel. Studies which examined the effectiveness and/or efficacy of wearing face coverings or factors related to adherence to face covering use were excluded. Published research and pre-publication articles were included. Reviews, position/discussion papers, conference abstracts, protocol papers, modelling studies, case reports and studies published in languages other than English were excluded (see Table 1).

**Table 1.** Inclusion criteria.

| Inclusion Criteria | Exclusion Criteria |
| --- | --- |
| • Papers published from 1 January 2020 to present day at time of search (17<sup>th</sup> May 2022)<br>• All populations and all public settings other than healthcare settings<br>• Quantitative or qualitative research relating to the impact of face covering use on adherence to other protective behaviours during the COVID-19 pandemic<br>• Face covering use: *i.* All types of face covering, including (but not limited to) handmade and commercial cloth coverings (cloth, cotton, gauze, etc), and medical coverings such as surgical face masks. *ii.* Face covering mandate.<br>• Experimental studies, observational studies, qualitative studies, laboratory studies, any other primary data, secondary data analyses.<br>• Published research and pre-publication articles | • Healthcare settings<br>• Papers not in English<br>• Systematic reviews, narrative reviews, guidelines, position/discussion papers, conference abstracts, protocol papers, modelling studies and case reports.<br>• Studies which examined the effectiveness and/or efficacy of wearing face coverings or factors related to adherence to face covering use |

### 2.2 Search strategy

A systematic search was conducted by UKHSA researchers on 17^th^ May 2022 for papers from January 2020 until the date of searches. Sources searched included Ovid Medline, Ovid Embase, Ovid PsychInfo, Ovid EmCare, medRxiv preprints, Research Square and WHO COVID-19 Research Database. Search terms included terms related to COVID-19 (e.g. COVID-19, coronavirus, Sars-COV2), face coverings (e.g. mask, face cover, mouth covers) and protective behaviours (e.g., social distancing, handwashing, face-touching). A complete list of search terms is available in S1 Appendix. Forward and citation searches of included papers were also conducted on September 16^th^ 2022, and eligible papers from January 1 2020 to that date were also included.

### 2.3 Study identification

Selection and screening of papers followed a systematic search method following a Preferred Reporting Items for Systematic Reviews and Meta-Analyses (PRISMA) framework (10). The study identification process is detailed in Figure 1 and contains full details of studies included and excluded at each stage. In the first stage of title and abstract screening, 10% of records were assessed in duplicate by two reviewers, and disagreements were resolved by discussion and consensus. In the second stage of title and abstract screening one reviewer assessed the remaining 90% of records. The screening tool Rayyan (11) was used for both these first and second stages. All relevant records were then screened by full text by one reviewer and checked by a second.There were five reasons for exclusion: (i) The direction of analysis was incorrect e.g., the study investigated behavioural predictors of face-covering use rather than vice versa (n = 5), (ii) the indepdent variables or outcome variables were not relevant to the research question (n = 14), (iii) the study did not comprise primary empirical research (n = 6), (iv) the paper was a previously undetected duplicate (n = 5), and (v) the paper was not written in English (n = 1). By the end of this process, 47 papers were deemed eligible and included in the review.

**Fig. 1.**
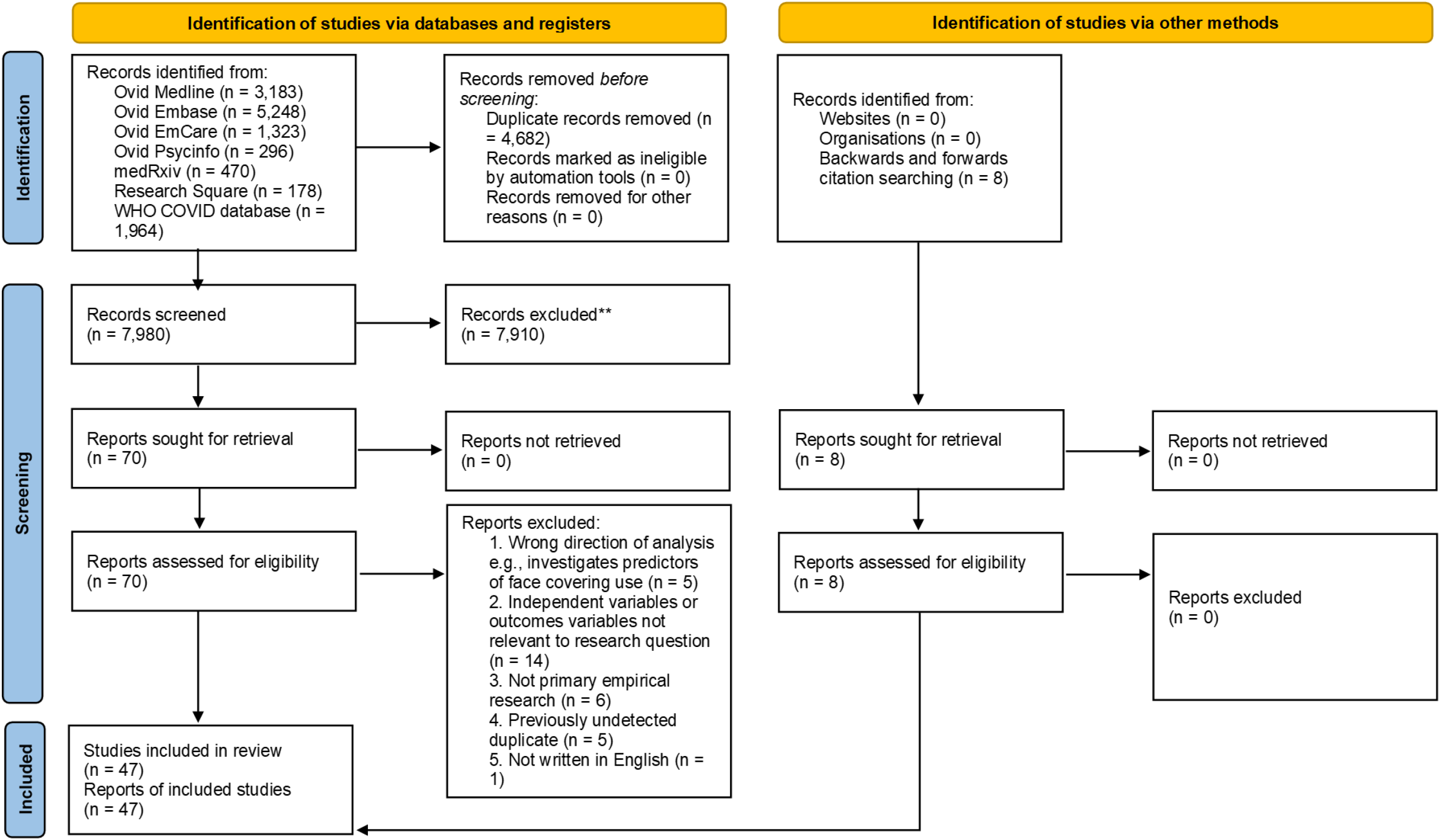
PRISMA 2020 flow diagram of the identification of studies process.

### 2.4 Data extraction and synthesis

Data was sought for any relevant COVID-19 protective behaviour outcomes; at the completition of extraction, these comprised the following behavioural outcomes: physicial distancing, mobility, face-touching, hand hygiene, close contacts and generalised measures of protective behaviour. All results that were compatible with each of these outcomes were included.The following information was then collated for each study: country, publication status, population and sample size, methods, key outcome variables and main results. This was initially collated into one table which included an outcome variable column. This table was then divided by outcome variable into six smaller data extraction tables, and a narrative summary of results was produced, structured by these six outcome variable categories.

Data extraction was completed for each included study by one reviewer and independently checked by a second reviewer, with discrepancies resolved by discussion. Any evidence that was not directly relevant to the review question was not extracted.

### 2.5 Quality assessment

Quality including risk of bias was evaluated by one reviewer using the Quality Criteria Checklist (QCC) tool which can be used to assess the methodological quality of a study (12). Using the QCC, studies were characterised as high, medium or low quality.

This checklist tool is composed of 10 questions, four of which are considered critical (questions on selection bias, group comparability, description of exposure/assessment of transmission routes, and validity of outcome measurements); full details can be seen in S2 Appendix. A study was rated as high methodological quality if the answers were yes to the four critical questions plus at least one of the remaining questions. A study was rated as low methodological quality if answers were no to >50% of the 10 questions. Otherwise, the study was rated as medium methodological quality. In line with a systematic review which found that self-report methods of capturing adherence to protective behaviours can over-estimate objectively observed adherence by up to a factor of five (13), any studies which employed a self-report methodology were deemed not to satisfy the validty of outcome criterion, and so were necessarily limited to a quality score of medium or below. An additional reviewer independently assessed five of the studies, and conflicting findings were discussed. Following discussions, a potential source of bias was identified in the first reviewer’s application of one of the tool’s questions, and all studies were subsequently re-evaluated to correct for this bias.

## 3. Results

### 3.1 Overview of search results

The initial searches generated 12,662 articles. After de-duplication, 7,980 records remained, and title and abstract screening resulted in 70 papers being accepted for full text screening. Following full text screening, 39 papers were accepted for inclusion in the review. Backwards and forwards citation searches were conducted which yielded a further 8 papers for inclusion, meaning the total number of papers included in the review was 47. Quality appraisal of the articles revealed that nine were of a high quality, 33 of a medium quality, and five of a low quality. Finally, of the papers included nine were pre-prints, 35 were peer-reviewed publications, and three were published conference papers.

Note that in the paragraph that follows, frequency of studies are described for each of several categories, e.g., methodology employed, face-covering intervention investigated, protective behaviour investigated, and means of measurement employed (objective or self-report). The total of the n’s in each category always exceeds the total number of papers included (47). This is because some papers included more than one study, more than one methodology, or were concerned with more than one face-covering intervention or protective behaviour.

All studies included used quantitative methods, and none used qualitative methods. These included field experiments (n = 7), natural experiments (n = 14), lab experiments (n = 14), observational studies (n = 10) and cross-sectional studies (n = 9). Of these, 30 studies measured behaviour through observation, and 22 measured behaviour through self-report. Of the self-report studies, 10 were concerned with current or previous behaviour, and 12 with expected or intended behaviour. Though all included papers necessarily investigated the effect of face covering use on protective behaviours, the exact intervention of focus in each case was varied and fit into one of 5 categories: face covering use (self) i.e., the act of wearing a face covering oneself (n = 21); face covering use (other) i.e., in some way interacting with someone else wearing a face covering (n = 18); face covering use (self and other) i.e., face covering wearing on behalf of oneself and another person with whom one is interacting or is in proximity to (n = 1); face covering policy i.e., the implementation of a face covering mandate (n = 19); and face covering intervention package i.e., a community programme designed to increase uptake of face covering usage (n = 1). The protective behaviour(s) in each case also varied, and in each case fit into one of 6 categories: physical distancing i.e., interpersonal distance (n = 30); mobility i.e., the extent to which people leave their home and visit public or residential spaces (n = 13); face-touching (n = 7); hand hygiene e.g., hand washing, avoidance of handshakes (n = 5); close contacts i.e., the extent to which people have close and extended interactions with people outside of their household (n = 4); and generalised protective behaviour i.e., a single measure to encapsulate two or more COVID-19 protective behaviours other than face-covering use, e.g, hand-washing, physical distancing and staying at home (n = 2). Full details on each paper can be seen in the data extraction tables in S3 Appendix.

Some studies initially appeared to meet the inclusion criteria but were ultimately excluded. For example, one study investigated how face covering use affected *perceptions* of physical distance between oneself and another person, however, since it did not investigate how face covering use affected distancing *behaviour*, it was not deemed to be eligible for inclusion (14). Similarly, another study investigated participants’ judgements on how face covering use moderates the relationship between a given physical distance and transmission risk was not included since, although its focus was related to the review’s objectives, it did not directly address it, in that it did not investigate how face covering use directly impacted adherence to protective behaviours (15).

The section below presents findings in relation to each of these six behaviours and describes the extracted data in terms of the effect that face covering use has on each type of behaviour, also exploring any patterns with regards to how the effect of face covering use varies by the type of intervention, and the type of study design.

### 3.2 Physical distancing (see table 2a in S3 Appendix for full details)

Outcomes were mixed and varied depending on the type of study. Findings from the majority of lab experiments (10 of 13) indicated that physical distancing was lower in conditions where a face covering was used by another person compared with conditions where it was not (16-23), (24: two studies), and two of these studies also found that physical distancing was lower when a face covering was used by the self (24: two studies). Additionally, one of these studies investigated how physical distancing in conditions with and without face covering use compared with a baseline measure (that is, physical distancing outside the context of face covering use) (16). It was found that physical distancing was significantly greater than the baseline in conditions without face covering use, but not significantly different in conditions with face covering use, potentially suggesting that differences between conditions is driven by participants moving *away* from people without face coverings (relative to where they would normally position themselves), rather than *towards* people with face coverings. Of the remaining three studies, one found that physical distancing was lower in conditions of face covering use (other), but that this effect was conditional on situational factors such as effort required to maintain physical distance, and age of the other person (25), and two found that distancing was greater in the case of face covering use (other) (26), (27: Study 2).

In contrast, the majority of field experiments (5 of 6) found that physical distancing was either greater or not significantly different in conditions where face coverings were used, compared to conditions where they were not. Two studies found that physical distancing was greater in conditions where face coverings (self or other) were used compared to when they were not (28), (27: Study 1), with a third study finding that physical distancing was greater in areas where a face covering intervention package had been implemented compared to those where it had not (29). One study found mixed effects, finding that face covering use (self) had no effect on distancing, but that distancing was greater in conditions of face covering use (other) (30), while another study found no differences in distancing between conditions of face covering use (self and other) and conditions without face covering use (31). A minority of field experiments (1 of 6) found that physical distancing was lower in conditions of face covering use (other), but that this effect was conditional on gender of participant, race and social status of confederate, and face covering policy implemented at the time (32). Additionally, two of the above studies also investigated whether the effect of face covering use on physical distancing differed between different time periods which varied by the face mask policy implemented at the time; one found that a distance-reducing effect of face covering use was stronger when a face covering use was mandated (but only for females) (32), whilst a second did not find a significant interaction between face mask policy and face covering use (30).

The four natural experiments explored the impact of face covering policy on physical distancing. Three of these found that physical distancing did not significantly differ according to whether face covering wearing was voluntary or mandatory (33: Study 2), (24: Study 2), (30), while a third found that the impact of face covering policy on physical distancing was dependent on context; attention given to physical distancing decreased when a face covering mandate was introduced on public transport, but increased when the mandate was expanded to bars and restaurants, and again when expanded to all public spaces (34).

The four observational studies found either a positive or a neutral relationship between face covering use or face covering policy and physical distancing. Three observational studies found no significant relationship between face covering use (self) or face covering policy and physical distancing (33: two studies), (35), while the fourth observational study found that physical distancing was greater in those who wore face coverings, and also that it was greater when a face covering mandate was implemented (36).

Four of the six correlational studies found that as self-reported face covering use (self or other) increased, so did self-reported physical distancing (37-40). One study showed mixed results for the relationship between face covering use and physical distancing, finding that those who reported wearing face coverings all the time were more likely to report adherence to physical distancing compared with those who reported never doing so, but those who reported wearing face coverings sometimes were more likely to report having reduced distancing with people than those who reported never doing so (41). The final correlational study found no relationship between face covering use (self) and physical distancing (42).

### 3.3 Mobility (see table 2b in S3 Appendix for full details)

Of the 10 natural experiments, 5 found that when face covering policies were in place mobility increased (i.e., people were less likely to stay at home) (43-47), 4 found that mobility was unchanged, (48-51) and one found that mobility decreased (i.e., people were more likely to stay at home) (52). The lab experiment found that mobility is likely to increase when policies are in place, as participants reported being more willing to use the London Underground when a face covering mandate was in operation (53).

Of the two cross-sectional studies, one found that self-reported adherence to personal protective behaviours, which included face covering use (self) predicted self-reported frequency of going out in public, and that this relationship was mediated by belief in ‘substitution myths’; the belief that one protective behaviour can be substituted for another, with risk staying constant (42). The second cross-sectional study found that those who reported always wearing a face covering outside their home were less likely to report visiting a friend, neighbour, bar or club (40).

### 3.4 Face-touching (see table 2c in S3 Appendix for full details)

The field experiment found that face covering use (self) resulted in less face-touching; there was a significant increase in the number of individuals who touched their eyes or hair when they were not wearing a face covering, and, when investigating areas not covered by a face covering (hair, forehead, eyes and ears) the absolute number of total touches was significantly higher in those not wearing one (54). Two of the six observational studies found similar results; those who wore face coverings were less likely to touch their face (55, 56). Two more observational studies found that results were dependent on the operationalisation of face-touching; when face-touching was defined as making contact with any part of the face (including touches to the mask), there was no significant relationship between face covering use (self) and face-touching, but when it was defined as making contact with specific parts of the face (not including touches to the mask), those who wore face coverings touched their faces less frequently (57: two studies). A minority of observational studies (one of 6) found a greater frequency of face-touching in those who wore coverings compared with those who did not (58). The final observational study found that face covering policy had no impact on face-touching; there was no statistically significant association between the face covering policy in operation and frequency of face-touching (59).

### 3.5 Hand hygiene (see table 2d in S3 Appendix for full details)

The natural experiment found that face covering policy had no impact on hand hygiene; attention paid to hygiene practices was not significantly different according to the implementation of different face covering policies (34). The three cross-sectional studies found that those who reported wearing face coverings more often were more likely to report also adhering to hand hygiene practices (37, 40, 41).

### 3.6 Close contacts (see table 2e in S3 Appendix for full details)

One of the two natural experiments found that number of close contacts was significantly lower when a face covering mandate was expanded from public transport to include restaurants and bars, and lower again when expanded to all public spaces (34). The other natural experiment found that a face covering mandate did not affect whether participants cancelled or postponed personal or social activities (51).

One of the three cross-sectional studies found that the relationship between face covering use (self) and number of close contacts was mixed; those who reported wearing face coverings all the time were more likely to report spending less than 15 minutes in close contact with someone compared with those who reported never wearing face coverings, while those who reported sometimes wearing face coverings were more likely to report spending over 60 minutes in close contact with someone compared with those who reported never doing so (41). The second cross-sectional study found that those who did not always wear face coverings outside work were more likely to have close contact with people they do not live with (40), whilst the final cross-sectional study found that self-reported face covering use outside of work significantly predicted a greater number of self-reported daily contacts (60).

### 3.7 Generalised protective behaviour (see table 2f in S3 Appendix for full details)

The term ‘generalised protective behaviour’ was used to categorise any studies whose outcome variable was a single measure which encapsulated two or more other COVID-19 protective behaviours besides face covering use e.g., hand washing, physical distancing and staying at home. Both studies found that self-reported face covering use (self) was associated with self-reported behaviours that are likely to reduce the spread of COVID-19. In one study the majority of participants reported that their adherence to other protective behaviours was either unchanged or greater when they wore a face covering compared with when they did not (61), while the other study found that self-reported face covering use (self) was associated with self-reported reduced engagement with COVID-19 related risky behaviour (62).

## 4. Discussion

Although face coverings have been found to reduce the transmission of airborne diseases, concerns have been raised as to the effect that their use may have on adherence to other COVID-19 protective behaviours. While several studies which explore this issue have been conducted, they had not previously been brought together in a review. This systematic review has identified and summarised the findings of 47 papers which collectively investigate the effect of face covering use on other COVID-19-related protective behaviours, namely, physical distancing, mobility, face-touching, hand hygiene, close contacts and generalised protective behaviour. Findings showed that results varied considerably, and that the nature of the variation often depended upon the specific behaviour investigated and the type of study employed to investigate it. The sections below, organised by behaviour, summarise and discuss the findings.

### 4.1 Physical distancing

One of the clearest patterns emerging from the studies relating to physical distancing was the distinction between the respective findings of lab experiments and field experiments. The majority of lab experiments found that physical distancing was lower in conditions of face covering use (compared with conditions without face covering use), whereas the majority of field experiments found that physical distancing was either greater or unchanged in conditions of face covering use.

A number of considerations should be made when interpreting these results, starting with the respective merits of field and lab experiments. There are three main advantages of lab experiments. First, the majority of the lab experiments employed a within-subjects design, meaning that individual differences which may impact on physical distancing (e.g., age, gender, risk perception, conscientiousness) are essentially controlled for in a way that cannot be done with field experiments (although estimations of such variables e.g., age was in some cases recorded and controlled for in the field experiments). Second, lab experiments allow for more precision in data collection. The lab experiments typically used an on-screen slider for participants to indicate their preferred distances, or a virtual reality environment in which computers captured the distances participants maintained from virtual agents. In contrast, some of the field experiments employed human judgement to determine whether a physical distancing guideline had been violated, increasing the risk of bias. However, four of the six field experiments used some form of electronic sensor to collect data (all of which found that physical distancing was greater or unchanged in conditions of face covering use). Third, lab experiments are able to examine the impact of face covering use on physical distancing in a context which is agnostic of face covering policy (mandatory vs voluntary), in contrast to field experiments which are necessarily conducted under periods of either mandatory or voluntary face covering wear. The impact of face covering use on other protective behaviours may be related to whether face covering use is mandatory or voluntary. For example, during periods of voluntary face covering wear the wearing of a face covering may act as a social cue to keep a greater distance (8); however, face covering wear may not act as the same social cue during periods of mandatory face covering wear (since all will be wearing face coverings). The ability to examine behaviour in a context which is agnostic of face covering policy may therefore be an advantage.

However, whilst field experiments are less controlled than lab experiments, they are more ecologically valid. A key advantage of field experiments compared to lab experiments is that the disease transmission risk is real, not hypothetical; therefore a person’s approach to physical distancing has a real impact on their own risk and on others’ risk of contracting and spreading COVID-19. In all but one of the six field experiments not only were participants blinded to the experimental condition they were assigned to, but they were also unaware that they were taking part in an experiment at all, and so responses were unlikely to biased by, for example, demand characteristics. Behaviour captured in field experiments may therefore be more representative of and generalisable to wider populations than that collected during lab experiments, in which participants could second-guess research objectives, potentially causing demand characteristics. Additionally, in the majority of lab experiments, rather than being measured for actual behaviour, participants were asked to indicate distances they would prefer from the avatars or characters, allowing them to make a conscious decision which may or may not be indicative of how they might behave in real-life scenarios.

In addition to the lab and field experiments, the effect of face covering use on physical distancing was also investigated through a number of different methodologies; the majority of natural and observational studies tended not to find any significant associations between face covering use (or face covering policy) and physical distancing, and cross-sectional studies tended to find positive associations between self-reported face covering-wearing and physical distancing. These different study types also have their respective strengths and weaknesses which should be considered when interpreting the results. The observational studies are necessarily high in ecological validity, however are less controlled. As such, the means of measurement (human judgement) may be lacking in precision and subject to bias, and furthermore, only claims of association (and not causality) can be made. For example it is likely that relationships between face covering-wearing and physical distancing are driven by confounding variables (e.g., risk perception, age, conscientiousness) impacting on both behaviours. The natural experiments, though again high in validity, are types of observational studies and so are subject to the same qualifications. Cross-sectional studies are also unable to draw claims of causality due to the likely impact of confounding variables, and may also be limited in their precision since measures are self-reported and not necessarily indicative of real-life behaviour.

A final consideration that should be made when interpreting the results is to consider the nature of the intervention in each case. Specifically, consideration should be given to: whether the independent variable (IV) is face covering condition of the self, face covering condition of another person, face covering policy, or some combination of any of the three; and the way in which physical distancing is operationalised in each case. Whilst there is no clear pattern with regards to how these different interventions and operationalisations of distancing impact outcomes, it is important to consider such differences when interpreting findings. Overall, it is clear that there is no consensus in relation to the impact of face covering use on physical distancing and that further research is required to bring the merits of different study types together and capture more reliable, and perhaps more consistent findings with regards to the effect of face covering use on physical distancing.

### 4.2 Mobility

The findings on mobility are also inconclusive, although there is more evidence to suggest that face covering policies increase mobility than there is to suggest that they decrease mobility. Natural experiments tended to compare changes in mobility from a baseline between areas and periods of time that varied according to the face covering policy implemented. The findings arising from such studies are therefore likely to be confounded by any number of other variables which are likely to impact upon mobility (e.g., other COVID-19 public health measures, rate of infection at time/area of data collection) and claims of causality must be made with caution. Generally speaking, however, these studies employ rich data sources and complex statistical models to account for these variables as far as is possible, and so conclusions made from these studies can be arrived at with a certain level of confidence. If it is the case that implementing face covering policies increases mobility, a plausible explanation may be that a mandate instils confidence in people that if they are to leave home others are more likely to wear a face covering, leading to a greater sense of security. Further qualitative work could be conducted to investigate how face covering policies impact upon people’s decision to leave home and move around their communities.

Two cross-sectional self-report studies which investigated how face covering use by the self impacted on mobility were inconclusive, with relationships found in both directions (40, 42). Interestingly however, in one case it was found that a positive relationship between self-reported adherence to personal protective behaviour (including wearing a face covering) and going out in public with people outside ones’ household was mediated by a belief that one protective behaviour can be safely substituted for another (42), providing evidence in favour of the risk compensation hypothesis.

### 4.3 Face-touching

The impact of face covering use on face-touching was the most consistent of all the behavioural outcomes, with a general trend for face-touching to be lower in conditions of face covering use than in conditions without. This was found in a field experiment (54) and four of six observational studies investigating the relationship between face covering use by the self and face-touching (55, 56), (57: two studies). Whilst observational studies are subject to the same caveats described in the physical distancing subsection the general pattern of a negative relationship between face touching and face covering use suggests that even if risk compensation is at play, it is not strong enough to result in a positive relationship. Furthermore, that this pattern is corroborated by the experimental study boosts the claim that face covering use might reduce face touching behaviour. It should however be noted that different operationalisations of face-touching were employed across these studies, and that in cases where significant effects or associations were found, they did not always apply to all parts of the face. Further research using experimental designs is required in this area, but on the basis of the studies reviewed to date, it seems reasonable to conclude that face covering use reduces instances of face-touching.

### 4.4 Hand hygiene

Across four studies (one natural experiment and three cross-sectional studies), three studies found a positive relationship between either face covering use or face covering policy and hand hygiene practices and a fourth found no significant relationship. The three studies which found positive relationships were cross-sectional (37, 40, 41) and based on self-report surveys; findings therefore show that those who report wearing face coverings are more likely to also report practicing good hand hygiene, but the cross-sectional nature of the studies means a causal relationship cannot be established. Furthermore, these studies can only inform us on how the isolated practices of wearing a face covering and practising hand hygiene relate to one another and cannot tell us whether hand hygiene is affected *whilst wearing* a face covering. The natural experiment (34), which employed face covering policies as an IV and found no significant effect on self-reported attention paid to hygiene practices may be more informative, suggesting that implementing a face covering mandate has no ‘collateral’ effect on hand hygiene practices. It should be noted however that this study is also subject to the caveats around accuracy associated with self-report data. In summary, the literature on the effect of face covering use on hand hygiene is limited and relies on self-report data, possibly because hand hygiene is a relatively private practice that is difficult to investigate using an experimental or observational design. On the basis of the literature reviewed however, it can be said that there is no evidence thus far to suggest that face covering use causes a reduction in hand hygiene practices.

### 4.5 Close contacts

There was little consensus in the literature on close contacts. Across two natural experiments, it was found that face covering mandates either had no effect on close contacts, or that they reduced them (34, 51). Across three cross-sectional studies the results were mixed with both positive and negative relationships found (40, 41, 60). It is therefore difficult to draw robust conclusions about the impact of face covering use on close contacts.

### 4.6 Generalised protective behaviour

Two cross-sectional studies (61, 62) investigated the relationship between face covering use by the self and generalised adherence to other protective behaviours and found a relationship between face covering use and increased adherence to other protective behaviours (or reduced undertaking of risky behaviours). As previously discussed, cross-sectional studies do not allow for claims of causality to be made, and confounding variables are likely to influence any relationships.

### 4.7 Limitations

This review is novel in drawing together current literature on the impact of face covering use on other protective behaviours during the COVID-19 pandemic, and contributing to an understanding of risk compensation in this context. However, some limitations should be acknowledged when considering this review’s findings. First, despite every attempt to apply the inclusion and exclusion criteria systematically and objectively, it is likely that some level of subjectivity may have impacted on paper screening and selection. To mitigate against this, 10% of the title and abstract screening was undertaken by two researchers, and any disagreements resolved by discussion and consensus. The full-text screening was also checked by a second researcher. Second, due to resource constraints (namely, the linguistic limitations of the researchers), only papers published in English were included in the review, inevitably creating a geographical bias. For example, 32 of the 47 papers described studies carried out in the UK, Western Europe, Canada, North America or Australia.

### 4.8 Conclusion

Overall, findings relating to the impact of face covering use or policy on other protective behaviours are inconsistent, varying both according to the particular behaviour in question, and by study type. Evidence relating to the potential impact of risk compensation is also inconsistent; while some studies suggest risk compensation may play a role in shaping behaviour, others suggest it has no impact. Findings are particularly inconsistent in relation to physical distancing, with considerable discrepancy found between study types. Whilst assessments of behaviour in real-life settings tend to find that face covering use either increases or has no impact upon physical distancing, lab experiments tend to suggest that physical distancing decreases in conditions of face covering use. There is some suggestion that the impact of face covering use on physical distancing could be affected by whether or not mandatory face covering policies are in effect (which would indicate that conclusions drawn from a voluntary context should not be directly applied to a mandatory context and vice versa) but findings in this area are mixed and further assessment of this moderating effect is warranted. Of the other behaviours, findings (though mixed) tend to suggest that face covering mandates may increase mobility and that face covering use may decrease face touching. While some studies suggest a relationship between face covering use and increased hand hygiene practices and adherence to other protective behaviours, these studies are predominately cross-sectional and rely on self-report measures, and do not allow conclusions to be drawn about the impact of face covering use on adherence to these behaviours. Findings relating to the impact of face covering use on close contacts is mixed, and further research is needed.

Overall, this review highlights that there is no clear consensus in the literature as to whether face covering use improves adherence to other protective behaviours, reduces adherence to other protective behaviours, or has no impact. There are two key recommendations which can be made from this review: 1) further research, using more robust research designs, is needed to establish whether face covering use affects different types of protective behaviours; and 2) any policy decisions made in relation to recommended or mandatory face covering use must take into account the inconsistencies and caveats in the evidence base, recognising that current evidence does not allow firm conclusions to be drawn about the potential impact of face covering use on adherence to other protective behaviours. Recommendations for future research are summarised below.

#### 4.8.1 Recommendations for future research

- Further field experiments (or more ecologically valid lab experiments) investigating the effect of face covering use on physical distancing. Such studies might make a particular focus of investigating:

- Differing effects of face covering use by the self and face covering use by another person
- How different face covering policies might moderate the relationship between face covering use and physical distancing. For example, a field experiment could be conducted whereby two manipulations occur; one whereby the setting varies according to the face covering policy implemented (mandatory or voluntary), and one whereby a confederate appears either wearing or not wearing a face covering. Physical distancing from the confederate would then be measured. This would allow for the direct comparison between the moderating effects of voluntary and mandatory settings with more control over extraneous factors.
- How effects might vary according to the operationalisation of distancing; for example, whether the absolute distance is measured, or whether a binary variable capturing whether or not a particular distance or guideline is adhered to is captured
- Lab experiments replicating those cited in this review, but which vary by the framing of physical distancing (i.e., self-protective or prosocial)
- Qualitative research investigating how differing face covering policies affect attitudes towards leaving the home and moving around the community
- Experimental studies investigating the effect of face covering use and face covering policies on face-touching, hand hygiene and close contacts

## Data Availability

All relevant data are within the manuscript and its Supporting Information files.

## 5. Acknowledgements

The authors thank Dr Joanna Milward for contributing to the development of the protocol and search terms, Nicola Pearce-Smith for creating the search strategies and running the searches themselves, as well as providing guidance on screening, and Megan Phillips for contributing to the quality appraisal as a second reviewer.

## 7. Supporting information captions

**S1 Appendix. Search strategies**

**S2 Appendix. Quality criteria checklist**

**S3 Appendix. Data extraction tables**

**S4 Appendix. PRISMA 2020 checklist**

**S5 Appendix. PRISMA 2020 abstract checklist**

**S6 Appendix. Protocol**

