## Supplementary material for "Effect of face-covering use on adherence to other COVID-19 protective behaviours: a systematic review": S2 Appendix

### Quality Criteria Checklist: Primary Research

#### Symbols Used

+ **Positive:** Indicates that the report has clearly addressed issues of inclusion/exclusion, bias, generalizability, and data collection and analysis.

-- **Negative:** Indicates that these issues have not been adequately addressed.

∅ **Neutral:** Indicates that the report is neither exceptionally strong nor exceptionally weak.

#### *Quality Criteria Checklist*: Primary Research

| RELEVANCE QUESTIONS | |
|---|---|
| 1. Would implementing the studied intervention or procedure (if found successful) result in improved outcomes for the patients/clients/population group? (NA for some Epi studies) | Yes No Unclear N/A |
| 2. Did the authors study an outcome (dependent variable) or topic that the patients/clients/population group would care about? | Yes No Unclear N/A |
| 3. Is the focus of the intervention or procedure (independent variable) or topic of study a common issue of concern to dietetics practice? | Yes No Unclear N/A |
| 4. Is the intervention or procedure feasible? (NA for some epidemiological studies) | Yes No Unclear N/A |
| ***If the answers to all of the above relevance questions are "Yes," the report is eligible for designation with a plus (+) on the Evidence Quality Worksheet, depending on answers to the following validity questions.*** | |
| VALIDITY QUESTIONS | |
| **1. Was the <u>research question</u> clearly stated?**<br>1.1 Was the specific intervention(s) or procedure (independent variable(s)) identified?<br>1.2 Was the outcome(s) (dependent variable(s)) clearly indicated?<br>1.3 Were the target population and setting specified? | Yes No Unclear N/A |
| **2. Was the <u>selection</u> of study subjects/patients free from bias?**<br>2.1 Were inclusion/exclusion criteria specified (e.g., risk, point in disease progression, diagnostic or prognosis criteria), and with sufficient detail and without omitting criteria critical to the study?<br>2.2 Were criteria applied equally to all study groups?<br>2.3 Were health, demographics, and other characteristics of subjects described?<br>2.4 Were the subjects/patients a representative sample of the relevant population? | Yes No Unclear N/A |
| **3. Were <u>study groups comparable</u>?**<br>3.1 Was the method of assigning subjects/patients to groups described and unbiased? (Method of randomization identified if RCT)<br>3.2 Were distribution of disease status, prognostic factors, and other factors (e.g., demographics) similar across study groups at baseline?<br>3.3 Were concurrent controls used? (Concurrent preferred over historical controls.)<br>3.4 If cohort study or cross-sectional study, were groups comparable on important confounding factors and/or were preexisting differences accounted for by using appropriate adjustments in statistical analysis?<br>3.5 If case control study, were potential confounding factors comparable for cases and controls? (If case series or trial with subjects serving as own control, this criterion is not applicable. Criterion may not be applicable in some cross-sectional studies.)<br>3.6 If diagnostic test, was there an independent blind comparison with an appropriate reference standard (e.g., "gold standard")? | Yes No Unclear N/A |
| **4. Was method of handling <u>withdrawals</u> described?**<br>4.1 Were follow up methods described and the same for all groups?<br>4.2 Was the number, characteristics of withdrawals (i.e., dropouts, lost to follow up, attrition rate) and/or response rate (cross-sectional studies) described for each group? (Follow up goal for a strong study is 80%.)<br>4.3 Were all enrolled subjects/patients (in the original sample) accounted for?<br>4.4 Were reasons for withdrawals similar across groups?<br>4.5 If diagnostic test, was decision to perform reference test not dependent on results of test under study? | Yes No Unclear N/A |
| **5. Was <u>blinding</u> used to prevent introduction of bias?**<br>5.1 In intervention study, were subjects, clinicians/practitioners, and investigators blinded to treatment group, as appropriate? | Yes No Unclear N/A |

| | |
|---|---|
| 5.2 Were data collectors blinded for outcomes assessment? (If outcome is measured using an objective test, such as a lab value, this criterion is assumed to be met.)<br>5.3 In cohort study or cross-sectional study, were measurements of outcomes and risk factors blinded?<br>5.4 In case control study, was case definition explicit and case ascertainment not influenced by exposure status?<br>5.5 In diagnostic study, were test results blinded to patient history and other test results? | |
| **6. Were intervention/therapeutic regimens/exposure factor or procedure and any comparison(s) described in detail? Were intervening factors described?**<br>6.1 In RCT or other intervention trial, were protocols described for all regimens studied?<br>6.2 n observational study, were interventions, study settings, and clinicians/provider described?<br>6.3 Was the intensity and duration of the intervention or exposure factor sufficient to produce a meaningful effect?<br>6.4 Was the amount of exposure and, if relevant, subject/patient compliance measured?<br>6.5 Were co-interventions (e.g., ancillary treatments, other therapies) described?<br>6.6 Were extra or unplanned treatments described?<br>6.7 Was the information for 6.4, 6.5, and 6.6 assessed the same way for all groups?<br>6.8 In diagnostic study, were details of test administration and replication sufficient? | Yes No Unclear N/A |
| **7. Were outcomes clearly defined and the measurements valid and reliable?**<br>7.1 Were primary and secondary endpoints described and relevant to the question?<br>7.2 Were nutrition measures appropriate to question and outcomes of concern?<br>7.3 Was the period of follow-up long enough for important outcome(s) to occur?<br>7.4 Were the observations and measurements based on standard, valid, and reliable data collection instruments/tests/procedures?<br>7.5 Was the measurement of effect at an appropriate level of precision?<br>7.6 Were other factors accounted for (measured) that could affect outcomes?<br>7.7 Were the measurements conducted consistently across groups? | Yes No Unclear N/A |
| **8. Was the statistical analysis appropriate for the study design and type of outcome indicators?**<br>8.1 Were statistical analyses adequately described the results reported appropriately?<br>8.2 Were correct statistical tests used and assumptions of test not violated?<br>8.3 Were statistics reported with levels of significance and/or confidence intervals?<br>8.4 Was “intent to treat” analysis of outcomes done (and as appropriate, was there an analysis of outcomes for those maximally exposed or a dose-response analysis)?<br>8.5 Were adequate adjustments made for effects of confounding factors that might have affected the outcomes (e.g., multivariate analyses)?<br>8.6 Was clinical significance as well as statistical significance reported?<br>8.7 If negative findings, was a power calculation reported to address type 2 error? | Yes No Unclear N/A |
| **9. Are conclusions supported by results with biases and limitations taken into consideration?**<br>9.1 Is there a discussion of findings?<br>9.2 Are biases and study limitations identified and discussed? | Yes No Unclear N/A |
| **10. Is bias due to study’s funding or sponsorship unlikely?**<br>10.1 Were sources of funding and investigators’ affiliations described?<br>10.2 Was there no apparent conflict of interest? | Yes No Unclear N/A |
| **MINUS/NEGATIVE (-)**<br>*If most (six or more) of the answers to the above validity questions are “No,” the report should be designated with a minus (-) symbol on the Evidence Worksheet.* | |
| **NEUTRAL (∅)**<br>*If the answers to validity criteria questions 2, 3, 6, and 7 do not indicate that the study is exceptionally strong, the report should be designated with a neutral (∅) symbol on the Evidence Worksheet.* | |
| **PLUS/POSITIVE (+)**<br>*If most of the answers to the above validity questions are “Yes” (including criteria 2, 3, 6, 7 and at least one additional “Yes”), the report should be designated with a plus symbol (+) on the Evidence Worksheet.* | |
