## Supplementary material for "Effect of face-covering use on adherence to other COVID-19 protective behaviours: a systematic review": S3 Appendix

S2 Appendix. Data extraction tables.

Table 2a - Physical distancing

| **Author, Year, Country, Quality and Publication Status** | **Population and Sample Size** | **Study Design and Procedure** | **Manipulation/**  **Comparison Groups** | **Key Outcome Variable(s)** | **Main Results** |
| --- | --- | --- | --- | --- | --- |
| Abaluck et al., 2021, Bangladesh  Quality: Medium  Preprint (not peer-reviewed) | General public  N = 64,937 adults in 600 villages | *Between-subjects experimental (field):* The effects of an intervention package to increase mask-wearing were tested for. Physical distancing was measured through direct observation at baseline and then once per week on weeks 1, 2, 4, 6, 8, and 10 after the intervention, 9am-7pm. | Intervention package implementation (implemented, not implemented)  Intervention included mask distribution and promotion, and role-modelling and advocacy by local leaders | Adherence to physical distancing (distance from nearest person at least one arm’s length) | Physical distancing significantly greater in villages where the mask interventions were implemented in comparison to paired control villages |
| Aranguren, 2022, France  Quality: Medium  Published | General public  N = 2722  First wave of data collection (June 2020) n = 1396  Second wave of data collection (September 2020)  n = 1326 | *Between subjects experimental (field):* A confederate approached pedestrians at traffic lights and asked for directions. A second experimenter observed and recorded data. A cylinder placed on top of a rolling suitcase was used as 1m reference for both confederate and observer. | (i) Confederate mask-wearing condition (ii) Pedestrian gender (iii) Confederate signalled social status (iv) Confederate race (v) Time of data collection (June - face mask use recommended, September - face mask use mandatory). | Adherence to physical distancing (minimum distance from confederate equal to or more than one metre) | No credible main effect of confederate mask wearing on distancing. Some simple effects:  (i) Males less likely to adhere when confederate wore a mask compared to when they did not; effect marginally greater in September and when the confederate signalled high social status.  (ii) In September, adherence-reducing effect of face masks amongst males stronger in those who interacted with a black confederate.  (iii) In mask-wearing conditions, females are less likely to adhere to physical distancing in September than they are in June. |
| Atzema et al., 2021, Canada and USA  Quality: Medium  Published | General public  N = 36,808 | *Observational:* Behaviour was observed by teams of data collectors 8am-9pm at indoor and outdoor spaces. | (i) Mask-wearing behaviour (ii) Face mask policy | ‘Breach’ (coming within 2m of another person, when both parties either had no mask or a mask that was worn incorrectly) | Significantly greater rate of distancing breaches in non-mask wearing group (including incorrect mask-wearing) than in correct mask-wearing group. Note that this finding is largely influenced by the fact that mask-wearing group included those who wore masks both correctly and incorrectly. According to the definition of of main outcome measure, it is impossible for those who are wearing masks to commit ‘a breach’, and this therefore ‘dilutes’ the rate of breaches in the mask-wearing group. By extension it also likely influences the secondary finding below.  Significantly lower rate of distancing breaches in mandatory settings than in voluntary settings |
| Betsch et al., 2020, Germany  Quality: Medium  Published | General public  N = 6,973 | *Cross-sectional:* Self-report survey including questions on behaviour and attitudes related to the COVID-19 pandemic | Mask-wearing behaviour | Physical distancing | Significant relationship between mask-wearing and physical distancing; individuals wearing masks often or always also reported greater adherence to physical distancing. |
| Biggio et al., 2022, Italy  Quality: Medium  Published | General public  N = 107  Main experiment  n = 82  Control experiment  n = 25 | *Within-subjects experimental (lab):* Two experiments were conducted in which participants were asked to indicate ‘reach distance’ and ‘comfort distance’ between an avatar representing themselves and an avatar representing another person. In addition, a survey took measures of variables such as trait anxiety, and whether the participant had been tested for COVID-19. | Main experiment: Avatar mask condition  Control experiment: No IV related to mask condition; all avatars presented without masks | Both experiments: Distance, operationalised as a function of the starting distance, the time taken for the participant to stop the avatars after they had begun the approach, and the total video duration. | Main experiment: Significant main effect of avatar mask condition on comfort distance; comfort distance was smaller when the avatar was not wearing a mask. Differences in comfort distance between avatar mask conditions correlated with trait anxiety.  Control experiment: Significant difference in comfort distance between avatar no mask condition in the main experiment, and the condition in the control experiment (where no avatars wore masks); comfort distance was significantly greater in the former. No significant difference in comfort distance between the avatar mask condition in the main experiment, and the condition in the control experiment (where no avatars wore masks). |
| Blanken et al., 2021, Netherlands  Quality: Medium  Published | General public (visitors of an art fair)  N = 639 | *Between-subjects experimental (field):* A number of behavioural interventions were implemented at an art fair, and data on distancing was collected via worn sensors | Intervention (Face masks required, no supplementary intervention) | Contacts (number of times a participant came within 1.5m of another participant) | No credible difference in contacts between face masks required condition and the no supplementary intervention condition |
| Bühler et al., 2021, Canada  Quality: Low  Preprint (not peer-reviewed) | General public  N = 11 | *Within-subjects experimental (lab):* Participants were set up in a virtual environment and asked to walk towards a target whilst avoiding collisions with any Virtual Reality Persons (VRP) in the scene. | VRP mask condition | (i) Minimum distance (ii) Maximum distance (iii) Onset distance (distance from VRP at which participant adjusts trajectory to avoid a collision) | Average onset distance significantly greater when VRP was wearing a face mask than when they were not  No significant differences in minimum or maximum distance |
| Cartaud et al. 2020, France  Quality: Medium  Published | General public  N = 457 | *Within subjects experimental (lab):* Participants were presented with a series of virtual characters appearing at various distances and asked to indicate whether they felt the distance was ‘appropriate’ or ‘inappropriate’. | Virtual character facial expression (masked, happy, angry, neutral) | Preferred interpersonal distance (IPD) from virtual character (average distance at which occurred the transition between ‘appropriate’ and ‘inappropriate’ responses) | Average preferred IPD significantly shorter when virtual characters wore a face mask compared to other conditions. |
| Doung-ngern et al., 2020, Thailand  Quality: Medium  Preprint (not peer-reviewed) | General public  N = 1050 | *Cross-sectional:* A phone interview was conducted which included questions on adherence to protective behaviours | Mask-wearing behaviour | Shortest distance of contact with others | Those who wore masks all the time more likely to maintain 1m+ physical distancing compared with those who never wear masks.  Those who wore masks sometimes more likely to have physical contact with others compared with those who never wear masks. |
| Griggs et al., 2021, USA  Quality: Medium  Published | General public   N = 613 | *Cross-sectional:* Self-report survey including questions on self-reported adherence to personal protective behaviours | Self-reported frequency of adherence to 11 personal protective behaviours, including mask-wearing | Ignoring social distancing (inverse of self-reported reduction in contact with others) | No direct effect of adherence to personal protective behaviours on ignoring social distancing |
| Gulbetekin et al, 2022, Turkey  Quality: Medium  Preprint (not peer-reviewed) | General public  N = 134 | *Within-subjects experimental (lab):* Participants were presented with a series of faces and asked to indicate on a slider their preferred distance from each person. | Mask condition of presented face | Social distance (mean preferred distance from each presented face) | Significantly greater preferred social distance from unmasked faces compared to masked faces |
| Iachini et al, 2020, Italy  Quality: Medium  Preprint (not peer-reviewed) | General public   N = 1293 | *Within-subjects experimental (lab):* Participants were asked to indicate on a Likert scale their preferred interpersonal space (IPS) from others | Hypothetical mask condition of another person | Preferred interpersonal space (IPS) | Preferred IPS significantly shorter in mask than in no mask condition |
| Jørgensen et al., 2021, Denmark  Quality: Medium  Published | General public  N = 106, 880 | *Natural experimental*: A self-report survey including questions on adherence to COVID-19 protective behaviours was conducted, and data compared between periods of different face mask policies. | Face mask policy | Distance attention (extent to which attention was given to physical distancing practices) | Public transport face mask mandate caused a decrease in distance attention, whilst expanding the mandate to bars and restaurants, and then to all public spaces both increased attention. |
| Kroczek et al, 2022, Germany  Quality: Medium  Published | General public  N = 84 | *Between- and within-subjects experimental (lab):* Participants were set up in a virtual supermarket environment and asked to make their way past a virtual agent to a target item and collect it. | (i) Virtual agent (VA) mask condition (within-subjects)  (ii) Trial type (direct path where keeping 1.5m distance required no additional effort, detour path where keeping 1.5m distance was effortful) (within-subjects) (iii) Virtual agent age (within-subjects) (iv) Participant mask condition (between subjects) | Interpersonal distance (IPD - the minimal distance between participant and VA) | Face masks worn by virtual agent significantly reduced IPD but only when participant did not wear a face mask. Further, this IPD-reducing effect was found only in detour path trials (where keeping 1.5m distance was effortful).  In direct path trial, agent face mask-use only reduced IPD towards younger agents (not older agents). In detour path trial, agent face mask use reduced IPD regardless of age. |
| Lama et al., 2022, USA  Quality: Medium  Published | General public  N = 1,804 | *Cross-sectional:* Self-report survey taking measures of variables related to COVID-19 public health measures | Perceived community mask-wearing | (i) Intention to get vaccinated, (ii) Intention to engage in social distancing | Perceived community mask-wearing significantly predicted both intentions to get vaccinated and to social distance; those who perceived that people in their community did not wear masks regularly reported lower intentions to get vaccinated and to socially distance |
| Lee & Chen, 2021, China and Taiwan  Quality: Medium  Published | General public  N = 200 | *Within and between subjects experimental (lab):* Participants were asked to position a virtual subject (avatar), or a virtual confederate, to a point where the distance between the two had just started to feel uncomfortable. | Confederate mask condition | Interpersonal space (IPS - distance at which participants indicated the distance had just started to feel uncomfortable) | Significant main effect of confederate mask condition; IPS greater when confederate was not wearing a mask compared to when they were. |
| Li & Colby, 2021, USA  Quality: Medium  Published | General public  N = 1,073 | *Cross-sectional:* Self-report survey taking measures of variables related to COVID-19 policies, perceptions and behaviours | (i) Actual face mask policy  (ii) Perceived face mask policy | Physical distancing (the extent to which participants maintained a 6-foot distance from others when visiting public spaces) | Perceived mask policy, not actual mask policy predicted social distancing; when people thought there was a face mask mandate, they were significantly more likely to adhere to physical distancing. |
| Liebst et al. 2022a,  Netherlands  Quality: Medium  Published | General public  Study 1  N = 383  Study 2  N = 423 | Study 1  *Observational:* Videos of general public were observed and data on mask-wearing and physical distancing were collected  Study 2  *Observational and natural experimental*: As above | Study 1  Mask-wearing behaviour  Study 2  (i) Mask-wearing behaviour (ii) Face mask policy | Studies 1 & 2  Adherence to physical distancing (maintaining 1.5m from strangers) | Studies 1 & 2  Nonsignificant association between mask-wearing and adherence to physical distancing  Study 2  No difference in adherence to physical distancing between areas with mandatory and voluntary face masking |
| Lisi et al, 2021, 28 Western countries  Quality: Medium  Published | General public  N = 242 | *Within-subjects experimental (lab):* Participants were presented with two avatars, one representing themselves, one representing a virtual confederate, and asked to indicate the minimum distance at which they would feel comfortable. | Other avatar protective equipment condition (wearing PPE; mask and gloves, not wearing PPE) | Interpersonal distance (IPD -distance indicated by the participant at which they would want the other avatar to stop while still being comfortable with that distance). | IPD significantly shorter when the avatar was wearing protective equipment compared to when they were not |
| Luckman et al., 2021, U.K.  Quality: Medium  Published | General public  Study 1, N = 401  Study 2, N = 400 | Study 1  *Within-subjects experimental (lab):* Participants were presented with digital scenarios which included avatars representing themselves and another person and asked to position their self-avatar at the closest distance they would keep from the other person in real-life.  Study 2  *Within-subjects experimental (lab) and natural experimental:* As above, with the addition of results being compared across different time periods varying by real-world face mask policy | Study 1  (i) Other-mask condition (ii) Self-mask condition  Study 2  As above, adding (iii)  face mask policy at time of data collection (voluntary, mandated on public transport) | Studies 1 & 2  Distance preference (the closest distance participants indicated they would keep from the stranger) | Significant main effects of both other-mask condition and self-mask condition; in both cases, distance preference was lower in the masked than the unmasked conditions.  No significant difference in distance preference between periods of different face mask policy. |
| Marchiori, 2020, Italy  Quality: Low  Preprint (not peer-reviewed) | General public   N = Unknown | *Between subjects experimental (field):* An ‘operator’ walked down Italian sidewalks with a ‘social distancing belt’; a belt with hidden sensor boxes which measures social distancing. | (i) Operator mask condition  (ii) Regional face mask policy in Veneto at the time of data collection | Social distancing (average distance between operator and others) | Social distancing significantly greater in all conditions where the operator wore some form of mask compared with when they did not.  None of these effects differ between different face mask policies. |
| Puttock et al., 2022, USA  Quality: Medium  Published | General public  N = 109,999 | *Observational:* Experimenters directly observed and assessed face mask use and distancing in public settings | Face mask policy | Adherence to social distancing (maintaining 6 feet from others) | Nonsignificant association between face mask policy and social distancing |
| Seres et al., 2021a, Germany  Quality: Medium  Published | General public  Study 1, N = 300  Study 2, N = 456 | Study 1  *Between-subjects experimental (field):* Experimenters joined a queue outside a shop and the distance that the next person joining the line stood from them was recorded using a light detection and ranging app on a mobile device.  Study 2  *Between-subjects experimental (lab):* Participants were presented with a picture of a person, were asked to imagine that that person was waiting in a queue outside a post office and were asked questions regarding distancing behaviour and their perceptions of the person. | Study 1  (i) Experimenter mask condition (ii) Participant mask-wearing behaviour  Study 2  Mask condition of pictured person | Study 1  (i) Distance (average distance kept by the participants from experimenter) (ii) Compliance (whether participants maintained the distancing mandate of 1.5m)  Study 2  (i) Estimated social distance of others (distance which the participant estimated someone would keep from the pictured person when joining the queue) (ii) Estimated preferred distance (estimation of the distance the pictured person would prefer the arriving person to keep) | Study 1  Distancing was greater in experimenter mask condition than experimenter no mask condition. The rate of compliance was also greater in the experimenter mask condition. Distancing was greater in the participant mask condition than the participant no mask condition  Study 2  Estimated social distance of others was not significantly different between mask conditions. Estimated preferred distance was significantly greater in mask condition than no mask condition. |
| Seres et al., 2021b, Germany  Quality: High  Published | General public  N = 480 | *Between-subjects experimental (field) and natural experimental:* Experimenters joined a queue outside a shop and the distance that the next person joining the line stood from them was recorded using a mobile augmented reality application. Results were compared across time periods varying by face mask policy. | (i) Experimenter mask condition (ii) Participant mask-wearing behaviour (iii) Face mask policy | Distance (distance kept by participants from experimenter) | Distance significantly larger in the experimenter mask condition than the experimenter no mask condition  No significant differences in distance according to participant mask-wearing behaviour  No significant differences in distance between face mask policies |
| Sheehan et al., 2020, USA  Quality: Medium  Published | Adult patients at a large integrated health system in Ohio who were active on an online health portal  N = 654 | *Cross-sectional:* Self-report survey including questions on face mask use, social distancing and hygiene behaviours in the previous seven days | Mask-wearing behaviour | Self-reported likelihood of avoiding others in public spaces and having close contact with people outside of household | Those who always wore masks outside their home were more likely to avoid others in public spaces. Those who did not always wear masks outside their home were more likely to visit a friend and have a close contact with people they did not live with. |
| Wang & Lee, 2022, China  Quality: Medium  Published (conference paper) | General public  N = 30 | *Within-subjects experimental (lab):* Participants were presented with two avatars, one representing themselves, one representing a virtual confederate, and asked to indicate preferred Interpersonal distance (IPD). | Confederate mask condition | IPD (distance indicated by the participant at which they would want the other avatar to stop while still being comfortable with that distance) | IPD significantly greater in the no mask than in the mask condition |
| Xiong et al., 2022, unsure  Quality: Medium  Published (conference paper) | General public   N = 31 | *Within-subjects experimental (lab):* Participants set up in a virtual environment and asked to indicate at which point they felt uncomfortable with the distance between themselves and a virtual confederate | Confederate mask condition | Interpersonal distance (IPD) (distance between the participant and the confederate at which the participant indicated they felt uncomfortable) | Significant main effect of confederate mask condition; IPD significantly greater when confederate did not wear a mask compared with then they did. |

Table 2b - Mobility

| **Author, Year, Country, Quality and Publication Status** | **Population and Sample Size** | **Study Design and Procedure** | **Manipulation/**  **Comparison Groups** | **Key Outcome Variable(s)** | **Main Results** |
| --- | --- | --- | --- | --- | --- |
| Bansal et al., 2022, U.K.  Quality: Medium  Published | General public  N = 961 | *Within-subjects experimental (lab):* A discrete choice experiment: Participants were presented with two scenarios based in the London underground which varied by a number of circumstantial factors and were asked to select in each case which situation they would prefer. | Face mask policy (as well as a number of other circumstantial factors not relevant for the purposes of this review) | Preferred scenario according to face mask policy | 70% of participants preferred scenarios where masks were compulsory. |
| Barros et al, 2022, 113 countries  Quality: High  Preprint (not peer-reviewed) | General public  N = Unknown | *Natural experimental:* Secondary data analysis of the effect of face mask policy on mobility using Google mobility data and the Worldwide Non-Pharmaceutical Interventions tracker for COVID-19. | Face mask policy | Mobility (percentage changes from a baseline in duration of time spent in residential areas, and in the movement in retail and recreation areas).  Country-level data used for all countries, except for US for which state-level data was used. | Face mask policies did not significantly impact on mobility |
| Chernozhukov et al., 2021, USA  Quality: High  Published | General public  N = Unknown | *Natural-experimental:* Secondary data analysis of the effect of face mask policy on mobility using Google mobility data and data on US state policies obtained from a database created by Raifman et al. (2020). | Face mask policy | Mobility (percentages changes from a baseline in visits and lengths of stay at four different public spaces e.g., transit stations, workplaces) | Very weak relationship between the mandate of face masks for business employees and mobility. In sum, face mask mandates do not impact on mobility. |
| Fracalossi de Moraes et al., 2022, Brazil  Quality: High  Published | General public  N = 6615 | *Natural-experimental:* Secondary data analysis of the effect of face mask policy on mobility using data provided by Brazilian geolocation company In Loco (later renamed Incognia). | Face mask policy | Social distancing (inverse of mobility-level according to data collected from In Loco) | Social distancing (inverse of mobility) significantly lower when both partial and full mask mandates were implemented, compared with no mandatory masking |
| Griggs et al., 2021, USA  Quality: Medium  Published | General public   N = 613 | *Cross-sectional:* Self-report survey including questions on self-reported adherence to personal protective behaviours | Self-reported frequency of adherence to 11 personal protective behaviours, including mask-wearing | (i) Going out in public (self-reported frequency of going out in public with people from outside their household) (ii) Hours spent away from home (self-reported number of hours in previous 2 weeks spent within 10 feet of non-household other people) | Significantly greater frequency of going out in public in those who adhere more to general protective behaviours; effect mediated by belief in substitution myths (beliefs that one protective behaviour can be substituted for another, and the level of risk stays constant)  No direct effect of adherence to personal protective behaviours on hours spent away from home. |
| Hosseini & Gittler, 2020, Europe and Northern America  Quality: Medium  Published (conference paper) | General public  N = 34 countries | *Natural experimental:* Secondary data analysis of the effect of face mask policy on mobility using Google mobility reports and extracted from the DELVE Global COVID-19 dataset. | Face mask policy | Workplace mobility and park mobility | Face mask policy significantly predicts greater workplace mobility.  It has the same relationship with park mobility, but the effect is much weaker. |
| Kovacs et al., 2020, Germany  Quality: Medium  Preprint (not peer-reviewed) | General public  N = Unknown | *Natural experimental:* Secondary data analysis of the effect of face mask policy on mobility using data from Google mobility reports. | Face mask policy | Mobility (percentage change in number of visits to, or time spent in, groceries and pharmacies, workplaces, and transit stations, relative to a baseline) | No significant effects of face mask policies on mobility. |
| Sheehan et al., 2020, USA  Quality: Medium  Published | Adult patients at a large integrated health system in Ohio who were active on an online health portal  N = 654 | *Cross-sectional:* Self-report survey including questions on face mask use, social distancing and hygiene behaviours in the previous seven days | Mask-wearing behaviour | Self-reported likelihood of visiting friend or neighbour’s residence, or a bar or a club | Those who did not always wear masks outside their home were more likely to visit a friend and have a close contact with people they did not live with. |
| Shin et al, 2021, USA  Quality: High  Published | General public  N = 7403 | *Natural-experimental:* Survey data was collected longitudinally between March 10 and June 23, 2020, asking questions on adherence to COVID-19 protective behaviours, and were analysed using a difference-in-difference model. | Face mask policy | (i) Avoiding public spaces (ii) Limiting eating at a restaurant (iii) Visiting a doctor | No significant differences in any of the mobility variables when face masks were mandated. |
| Wadud et al., 2022, Bangladesh  Quality: High  Published | General public  N = Unknown | *Natural experimental:* Secondary data analysis mobility levels between two time periods differing by face mask policy using Google mobility reports | Face mask policy | Mobility (number of visitors to six types of location in Bangladesh) | Level of mobility during a face mask mandate was significantly greater than the predicted values based on the previous period where there was no face mask mandate |
| Wang et al., 2022, 121 countries  Quality: High  Published | General public  N = 121 countries | *Natural experimental:* Secondary data analysis of the effect of face mask policy on mobility using data from Google mobility reports. | Face mask policy (unclear on different conditions – assumed mandatory vs. non-mandatory) | Mobility (percentage change in the number of visits and length of stay in public places, according to Google mobility reports) | Facial mask policies significantly predict greater mobility |
| Yan et al., 2021, USA  Quality: High  Published | General public  N = 48 USA states | *Natural experimental:* Secondary data analysis of the effect of state-level face mask policy on mobility using SafeGraph anonymized location data. | Face mask policy | (i) Time at home (device-weighted county mean of the median Census Block Group daily home dwell time in minutes)  (ii) Points of interest visitation (aggregation of each day’s number of visits to a particular site in a particular county) | Significantly decreased time at home and increased points of interest visitation following implementation of a face mask mandate |
| Zhao et al., 2022, USA  Quality: High  Preprint (not peer-reviewed) | General public  N = 3055 counties | *Natural experimental:* Secondary data analysis of the effect of face mask policy on mobility using SafeGraph anonymized location data. | Face mask policy | Mobility | Mobility is significantly greater when mask use is mandated |

Table 2c - Face-touching

| **Author, Year, Country, Quality and Publication Status** | **Population and Sample Size** | **Study Design and Procedure** | **Manipulation/**  **Comparison Groups** | **Key Outcome Variable(s)** | **Main Results** |
| --- | --- | --- | --- | --- | --- |
| Chen at al., 2020, China, Japan, South Korea, Western Europe, USA  Quality: Low  Published | General public  N = 7,580 | *Observational:* Videos recorded in public areas were searched for and the behaviour of those people present in videos was observed. | Mask-wearing behaviour | Face-touching behaviour (touching of the face with hands, cellular telephones, and other items, as well as eating) | Mask wearing was associated with a reduction in face-touching behaviours. |
| de Freitas et al., 2022, Brazil  Quality: Medium  Published | General public  N = 40 | *Within subjects experimental (field):* A 30-minute video interview was conducted. For half of its duration the participant wore a face mask, and for half they did not. | Face mask condition | Face-touching (Primary measure: number of participants who touched specified areas of their face at least once. Secondary measure: average number of touches to the specified face areas) | Number of individuals who touched their hair and eyes when not wearing a mask significantly greater than those who touched those areas when wearing a mask  Significantly greater number of touches to face areas in those not wearing a mask (but only when investigating touches to areas not covered by mask) |
| Kungurova et al., 2020, Saint Kitts  Quality: Low  Published | General public  N = 468 | *Observational:* Participants were observed in public spaces. It was recorded whether a mask was worn, and whether ‘errors’ wearing them were made (not covering nose, not covering the mouth, touching face). | Mask-wearing behaviour | Face-touching | A larger proportion of people wearing any face-covering (N95, surgical mask, manmade mask, bandana/scarf) displayed face-touching behaviour than those wearing no mask. |
| Liebst et al., 2022b, Netherlands  Quality: Medium  Published | General public   Study 1  N = 383  Study 2  N = 421 | Studies 1 & 2  *Observational:* Videos of general public were observed and data on mask-wearing and face-touching were collected. | Studies 1 & 2  Mask wearing behaviour | Studies 1 & 2  (i) Primary measure of face-touching (touching the face or mask at least once)  (ii) Three secondary measures of face-touching (touching of the face, mid face, and t-zone, but not the mask itself). | Studies 1 & 2  Nonsignificant association between mask condition and primary measure.  Significant negative association between mask wearing and secondary measure. |
| Shiraly et al., 2020, Iran  Quality: Medium  Published | General public  N = 1,000 | *Observational:* Participants in public spaces were observed for mask-wearing and face-touching behaviour | Mask-wearing behaviour | (i) Face-touching behaviour (number of touches to the face per hour) (i) Face-touching behaviour categorised by zone (mucosal, non muscoasl) | Face-touching (regardless of zone) significantly more frequent in those not wearing a mask, than those wearing a mask. Touching of the mucosal zone was significantly more frequent in those not wearing a mask. |
| Tao et al., 2020, China  Quality: Low  Published | General public  N = 61 | *Observational:* Bus surveillance videos were observed for face-touching behaviour | Face mask policy | Face-touching (number of times any area of the face was touched during a 30-minute interval) | Non-significant association between face mask policy and face-touching behaviour |

Table 2d - Hand hygiene

| **Author, Year, Country, Quality and Publication Status** | **Population and Sample Size** | **Study Design and Procedure** | **Manipulation/**  **Comparison Groups** | **Key Outcome Variable(s)** | **Main Results** |
| --- | --- | --- | --- | --- | --- |
| Betsch et al., 2020, Germany  Quality: Medium  Published | General public  N = 6,973 | *Cross-sectional:* Self-report survey including questions on behaviour and attitudes related to the COVID-19 pandemic | Mask-wearing behaviour | Hand hygiene (avoiding handshakes, washing hands) | Significant relationship between mask-wearing and hand hygiene; individuals wearing masks often or always also reported greater adherence to handwashing and the avoidance of handshakes |
| Doung-ngern et al., 2020, Thailand  Quality: Medium  Preprint (not peer-reviewed) | General public  N = 1050 | *Cross-sectional:* A phone interview was conducted which included questions on adherence to protective behaviours | Mask-wearing behaviour | Handwashing | Those who wore masks all the time or sometimes more likely to wash their hands often, compared with those who never wear masks. |
| Jørgensen et al., 2021, Denmark  Quality: Medium  Published | General public  N = 106, 880 | *Natural experimental*: A self-report survey including questions on adherence to COVID-19 protective behaviours was conducted, and data compared between periods of different face mask policies. | Face mask policy | Hygiene attention (extent to which attention was paid to hygiene practices e.g., washing hands, using hand sanitizer) | No effect on attention to hygiene across any of the policies. |
| Sheehan et al., 2020, USA  Quality: Medium  Published | Adult patients at a large integrated health system in Ohio who were active on an online health portal  N = 654 | *Cross-sectional:* Self-report survey including questions on face mask use, social distancing and hygiene behaviours in the previous seven days | Mask-wearing behaviour | Handwashing | Those who always wore masks outside their home washed hands more frequently than those who did not. |

Table 2e - Close contacts

| **Author, Year, Country, Quality and Publication Status** | **Population and Sample Size** | **Study Design and Procedure** | **Manipulation/**  **Comparison Groups** | **Key Outcome Variable(s)** | **Main Results** |
| --- | --- | --- | --- | --- | --- |
| Doung-ngern et al., 2020, Thailand  Quality: Medium  Preprint (not peer-reviewed) | General public  N = 1050 | *Cross-sectional:* A phone interview was conducted which included questions on adherence to protective behaviours | Mask-wearing behaviour | Duration of contact time with others within one metre | Those who wore masks all the time more likely to spend less than 15 minutes with others compared with those who never wear masks.  Those who wore masks sometimes more likely to spend over 60 minutes with others compared with those who never wear masks. |
| Jørgensen et al., 2021, Denmark  Quality: Medium  Published | General public  N = 106, 880 | *Natural experimental*: A self-report survey including questions on adherence to COVID-19 protective behaviours was conducted, and data compared between periods of different face mask policies. | Face mask policy | Contact behaviour (number of people participants had had close contact with) | Number of close contacts decreased with expansion of policy to restaurant and bars, and with the expansion to all public spaces, but not with the initial public transport policy change. |
| Sheehan et al., 2020, USA  Quality: Medium  Published | Adult patients at a large integrated health system in Ohio who were active on an online health portal  N = 654 | *Cross-sectional:* Self-report survey including questions on face mask use, social distancing and hygiene behaviours in the previous seven days | Mask-wearing behaviour | Self-reported likelihood of having close contact with people one does not live with | Those who did not always wear masks outside their home were more likely to have close contact with people they did not live with |
| Shin et al, 2021, USA  Quality: High  Published | General public  N = 7403 | *Natural-experimental:* Survey data was collected longitudinally between March 10 and June 23, 2020, asking questions on adherence to COVID-19 protective behaviours, and were analysed using a difference-in-difference model. | Face mask policy | Limiting social activity | No significant differences in limiting social activity when face masks were mandated |
| Van den Broek-Altenburg et al, 2021, USA  Quality: Medium  Published | Community members who had had an encounter with their primary care provider in the past 3 years  N = 1694 | *Cross-sectional:* Self-report survey including questions on variables such as work, living situations and behaviour. | Mask-wearing behaviour outside work | Number of daily contacts | Wearing a face mask outside of work significantly predicted a greater number of daily contacts with adults (but not older adults or children) |

Table 2f - Generalised protective behaviour

| **Author, Year, Country, Quality and Publication Status** | **Population and Sample Size** | **Study Design and Procedure** | **Manipulation/**  **Comparison Groups** | **Key Outcome Variable(s)** | **Main Results** |
| --- | --- | --- | --- | --- | --- |
| Guenther et al., 2021, U.K.  Quality: Medium  Published | General public  N = 1,254 | *Cross-sectional:* Self report survey including questions on real-world risk-taking behaviour in the context of COVID-19. | Mask wearing behaviour | Mask compensation index (average of self-reported scores to six questions about COVID-19 related behaviours when wearing a mask) | Participants significantly less likely to engage in COVID-19 related risky behaviours when wearing a mask. |
| MacIntyre et al., 2021, Australia, U.K., USA  Quality: Medium  Published | General public  N = 2343 | *Cross-sectional:* Self-report survey in which one question asked the extent to which participants changed their behaviour when wearing masks | Mask wearing behaviour | Self-reported changes in hand-washing and physical distancing when wearing a mask | 78.9% reported either no change or stricter adherence to other behaviours. |
