## Supplementary material for "Effect of face-covering use on adherence to other COVID-19 protective behaviours: a systematic review": S6 Appendix

Review methods were amended after registration. Please see the revision notes and previous versions for detail.

### Citation

Adam Millest, Holly Carter. Effect of face-covering use on adherence to other Covid-19 protective behaviours. PROSPERO 2022 CRD42022331961 Available from:
https://www.crd.york.ac.uk/prospero/display_record.php?ID=CRD42022331961

### Review question [1 change]

Does face-covering use affect adherence to other Covid-19 protective behaviours such as social distancing?

### Searches

Sources:

MEDLINE, Embase, medRxiv preprints, SSRN, PsycINFO, EmCare, WHO COVID-19 Research Database

Reference lists of relevant papers will also be searched, as well as relevant systematic reviews or evidence summaries identified.

Date of publication: 1 January 2020 to present.

Language: English.

Additional search strategy information can be found in the attached PDF document (link provided below).

### Types of study to be included

Included: Experimental studies, observational studies, qualitative studies, laboratory studies, any other primary data, secondary data analyses.

Excluded: Systematic or narrative reviews, guidelines, opinion pieces, modelling studies, case reports.

### Condition or domain being studied [1 change]

We are studying protective behaviours (e.g., social distancing, hand hygiene) within the context of the Covid-19 pandemic. These behaviours are intended to reduce transmission of the virus, and we are interested in the extent to which the use of a face-covering affects adherence to these behaviours.

### Participants/population

All populations.

### Intervention(s), exposure(s)

1. Face covering use: All types of face covering will be considered, including (but not limited to) handmade and commercial cloth masks (cloth, cotton, gauze, etc), and medical masks.

2. Face covering mandate.

### Comparator(s)/control

1. Lack of face covering use i.e. people not wearing a face covering.

2. Lack of face covering mandate.

### Context

Included: All public settings.

Excluded: Healthcare settings.

### Main outcome(s) [1 change]

Included: Adherence to other Covid-19 protective behaviours, including but not limited to:

- Social distancing

- Hygiene e.g., avoidance of face-touching, regular handwashing, etc

- Staying at home

- Reducing travel.

Excluded: Studies examining the effectiveness and/or efficacy of wearing face-coverings, studies examining factors related to adherence to face-covering use.

#### Measures of effect

Social distancing: Physical distance between participants/confederates/members of the public; number of times that a pre-determined conception of social distancing is violated; self-reported adherence to social distancing.

Hygiene: Number of observations of touching of the face; self-reported hygiene behaviours; self-reported adherence to social distancing.

Staying at home/reducing travel: Level of population-level mobility as measured through GPS data; self-reported adherence to stay-at-home guidelines.

### Additional outcome(s)

None.

### Data extraction (selection and coding)

Screening: Screening on title and abstract will be undertaken; at least 10% of titles will be assessed in duplicate by 2 reviewers, and the rest by 1 reviewer. Disagreements will be resolved by discussion and consensus.

Screening on full text will be undertaken by 1 reviewer and checked by a second.

Data Extraction: Summary information for each study will be extracted and reported in tabular form. Information will include country, setting, population, study design, outcomes measures, results and any relevant contextual data (such as local face mask recommendation at the time of the study).

This will be undertaken by 1 reviewer and checked by a second.

### Risk of bias (quality) assessment

Quality for quantitative studies will be assessed using the Academy of Nutrition and Dietetics quality criteria checklist (QCC) for primary research. This tool is not specific to nutrition and can be applied quickly to most study designs to consider core quality criteria and domains.

Quality for qualitative studies will be assessed using the Critical Appraisal Skills Programme (CASP) tool for qualitative studies.

Quality will be assessed by 1 reviewer and checked by a second.

### Strategy for data synthesis

A narrative synthesis will be provided. Following guidance from the ESRC methods programme we will develop a primary synthesis, explore the relationships in the data, and asses the robustness of the synthesis product.

### Analysis of subgroups or subsets

Variations across populations and subgroups, for example cultural variations or differences between ethnic, social or vulnerable groups will be considered, where evidence is available.

### Contact details for further information

Adam Millest

### Organisational affiliation of the review

Behavioural Science and Insights Unit (BSIU), UKHSA

### Review team members and their organisational affiliations

Mr Adam Millest. Behavioural Science and Insights Unit (BSIU), UKHSA

Dr Holly Carter. Behavioural Science and Insights Unit (BSIU), UKHSA

### Collaborators

Nicola Pearce-Smith. UKHSA

### Type and method of review

Epidemiologic, Narrative synthesis, Systematic review

### Anticipated or actual start date

22 May 2022

### Anticipated completion date [3 changes]

31 December 2022

### Funding sources/sponsors

The review will be funded by the UK Health Security Agency where we are employees

### Conflicts of interest

### Language

English

### Country

England

### Stage of review [1 change]

Review Ongoing

### Subject index terms status

Subject indexing assigned by CRD

### Subject index terms

Communicable Disease Control; Communicable Diseases; COVID-19; Hand Hygiene; Humans; Masks; Physical Distancing; Protective Factors; Public Health; Quarantine; SARS-CoV-2

### Date of registration in PROSPERO

18 May 2022

### Date of first submission

18 May 2022

### Stage of review at time of this submission [4 changes]

| Stage | Started | Completed |
|---|---|---|
| Preliminary searches | Yes | Yes |
| Piloting of the study selection process | Yes | Yes |
| Formal screening of search results against eligibility criteria | Yes | Yes |
| Data extraction | Yes | Yes |
| Risk of bias (quality) assessment | Yes | Yes |
| Data analysis | Yes | Yes |

### Revision note

I have updated the progress section to reflect the fact that data analysis and quality appraisal have been completed, and that we are finalising the draft for publication.

*The record owner confirms that the information they have supplied for this submission is accurate and complete and they understand that deliberate provision of inaccurate information or omission of data may be construed as scientific misconduct.*

*The record owner confirms that they will update the status of the review when it is completed and will add publication details in due course.*

### Versions

18 May 2022

20 May 2022

05 September 2022

21 September 2022

01 November 2022

03 January 2023
